# The potential effect of romosozumab on perioperative management for instrumentation surgery

**DOI:** 10.1101/2023.11.09.23298298

**Authors:** Koji Ishikawa, Soji Tani, Tomoaki Toyone, Koki Tsuchiya, Tomoko Towatari, Yusuke Oshita, Ryo Yamamura, Takashi Nagai, Toshiyuki Shirahata, Katsunori Inagaki, Yoshifumi Kudo

## Abstract

**Background:** Age-related changes in bone health increase the risk for complications in elderly patients undergoing orthopedic surgery. Osteoporosis is a key therapeutic target that needs to be addressed to ensure successful instrumentation surgery. The effectiveness of pharmacological interventions in orthopedic surgery, particularly the new drug romosozumab, is still unknown. We aim to evaluate the effect of 3-month romosozumab treatment on biomechanical parameters related to spinal instrumentation surgery, using the Quantitative Computed Tomography (QCT)-based Finite Element Method (FEM).

**Methods:** This open-labeled, prospective study included 81 patients aged 60 to 90 years, who met the osteoporosis criteria and were scheduled for either romosozumab or eldecalcitol treatment. Patients were assessed using blood samples, dual-energy absorptiometry (DXA), and QCT. Biomechanical parameters were evaluated using FEM at baseline and 3 months post-treatment. The primary endpoints were biomechanical parameters at 3 months, while secondary endpoints included changes in regional volumetric bone mineral density around the pedicle (P-vBMD) and vertebral body (V-vBMD).

**Results:** Romosozumab treatment led to significant gains in P-vBMD, and V-vBMD compared to eldecalcitol at 3 months. Notably, the romosozumab group showed greater improvements in all biomechanical parameters estimated by FEM at 3 months compared to the eldecalcitol group.

**Conclusion:** Romosozumab significantly increased the regional vBMD as well as biomechanical parameters, potentially offering clinical benefits in reducing post-operative complications in patients with osteoporosis undergoing orthopedic instrumentation surgery. This study highlights the novel advantages of romosozumab treatment and advocates further research on its effectiveness in perioperative management.

## Introduction

Recent medical advancements have significantly increased life expectancy, leading to a rise in the number of aged patients undergoing surgeries (1–3). The significant shift in the aging population highlights osteoporosis as an enormous problem in orthopedic surgery due to its impact on postoperative complications (4–6).

Osteoporosis, a global health concern, is not only an essential therapeutic target for extending healthspan, but also lifespan (7–9). Recent research has further investigated the links of osteoporosis with orthopedic surgery, aiming to optimize operative outcomes (10–13). Specifically, if instrumentation spinal surgery is indicated, adequate risk stratification, including bone health, is advocated to ensure optimal and safe outcomes, since bone fragility exacerbates postoperative complications such as loosening, junctional failure, and cage subsidence, potentially resulting in revision surgery. Indeed, previous studies have reported that 25-60% of patients with osteoporosis experienced postoperative complications, even following successful instrumentation surgery (14–16).

While dual-energy absorptiometry (DXA) is a reliable method for assessing bone health, earlier imaging studies highlight the heterogeneity of the bone mineral density (BMD) inside the vertebra, which is overlooked by DXA measurement (17). Additionally, recent findings indicate that regional volumetric bone mineral density (vBMD) around an implant, as measured by QCT, demonstrates stronger correlations with both intraoperative screw fixation and postoperative complications, including screw loosening and cage subsidence, compared to areal bone mineral density (aBMD) assessed by DXA. (6,18,19). Furthermore, Keaveney et al. demonstrated the superiority of CT-based biomechanical analysis over BMD testing alone in terms of predicting reoperation following spinal instrumentation surgery (5). Thus, a multidisciplinary approach including bone health assessment of the heterogeneity of the BMD in the vertebra as well as biomechanical analysis is recommended prior to surgery to an ensure optimal and safe outcome.

Pharmacologic therapy could play an important role in perioperative bone management for facilitating bone health, but its effectiveness for orthopedic implant surgery has not been systematically investigated (12,20). The possible advantage of osteoporosis treatments for instrumentation surgery, including teriparatide and bisphosphonate, specifically addressing issues like screw loosening and adjacent vertebral fractures, have been reported previously (18,20–23). Although the contribution of BMD to cage subsidence is described, to the best of our knowledge, there is no study examining the effects of osteoporosis treatment for cage subsidence (19). A significant challenge in biomechanical implant analysis in the clinical, particularly during pharmacological intervention, is the inability to conduct damage or destruction analysis. Our recent *in silico* biomechanical analysis using Finite Element Method (FEM) challenges these limitations, and showed that a 12-month course of denosumab treatment (an anti-RANKL antibody) is potentially beneficial for reducing postoperative screw loosening following spinal instrumentation surgery (18). This study lays the groundwork for understanding the impact of osteoporosis treatment on optimizing outcomes in instrumentation surgery. However, considering the onset of postoperative complications within the initial few months after surgery, there is a pressing need for treatments that show benefits quickly following administration (16).

Although increasing evidence suggests that osteoporosis treatment positively impacts orthopedic surgery, no standard approach exists for perioperative management. A substantial hurdle when incorporating osteoporosis treatments into surgical strategies is the extended time frame, typically around a year, required for the improvement of bone structure after treatment initiation. However, for patients requiring immediate surgery, delaying procedures for the benefits of osteoporosis treatment is not a viable option. This presents a challenge in determining the optimal timing and effectiveness of osteoporosis treatment for patients in need of immediate surgical intervention. Romosozumab may offer a solution to this issue, given that previous reports have shown its potential to increase BMD shortly after treatment administration in terms of both imaging and bone biopsy assessment (24–26).

Thus, we aim to evaluate the effects of a 3-month romosozumab treatment on spinal instrumentation surgery using QCT-based FEM. We have extended our earlier FEM methods to cover analysis for the risk of cage subsidence, a major complication that occur in 5-50% of cases following surgery and is linked with negative clinical outcomes (19). Here, we first demonstrated the potential benefits of short-term romosozumab treatment in improving bone health and reducing postoperative complications in patients with osteoporosis undergoing spinal instrumentation surgery.

## Methods

### Study subjects

This was an open-labeled prospective study of ambulatory patients 60 to 90 years of age who met the osteoporosis criteria (27). Between March 2019 and March 2021, 226 patients who were scheduled for Romosozumab (Romo) or Eldecalcitol (ELD) treatment, were assessed for inclusion, of which 81 patients participated in the present study.

The patients were divided into two groups based on their treatment (Romo: 69 patients, ELD: 12 patients). All patients received daily eldecalcitol (0.75 μg) and / or calcium (400-800 mg) except for four patients in the Romo group. These four exceptions were decided by physicians based on their baseline serum calcium levels to prevent hypercalcemia. The medication compliance related to eldecalcitol and calcium was assessed at each visit and it was confirmed that all patients consumed > 90% of the drugs over the course of the study. The study was approved by the local ethics committees of Yamanashi Red Cross Hospital and in accordance with the precepts of the Declaration of Helsinki. All patients provided informed consent before participation.

### Assessments

We measured the spine-areal BMD (spine-aBMD) using DXA (L1-4) (Hologic QDR series: Hologic, Waltham, MA) at baseline and 6 months. All DXA measurements were analyzed by a radiologist at a central site. The regional vBMD around the pedicle (P-vBMD) and vertebral body (V-vBMD) as well as biomechanical parameters were measured by QCT-based FEM at baseline and 3 months. The intra- and inter-observer coefficients of variation of BMD assessments have been previously described (6,18,28) The serum levels of albumin, calcium, and phosphorus as well as estimated glomerular filtration rate (eGFR) were evaluated at baseline. The bone turnover markers including serum levels of TRACP-5b and total-P1NP were assessed at baseline, then at 3 and 6 months following treatment. The primary endpoints were the biomechanical parameters at 3 months. Secondary endpoints included the changes in regional vBMD and bone turnover markers throughout the study.

### Three-dimensional volumetric bone mineral density

The details of the measurement of vBMD have been described in previous studies (6,18,29,30). CT data were acquired with a SOMATOM Definition AS+ multidetector-row CT scanner (Aquilion 16; Toshiba Medical Systems, Otawara, Japan) using predefined scanning conditions (x-ray energy, 120 kV; x-ray current, SD20; rotation speed, 0.5 s/rot; beam pitch, 0.95). For QCT scanning, a phantom (Mindways, Austin, TX, USA) was placed underneath the patients for BMD calibration, thereby ensuring measurement quality throughout the study. The vBMD at the vertebral body and pedicle (reference vertebra L4) was measured using MECHANICAL FINDER (Research Center of Computational Mechanics; version 10.0, Tokyo, Japan) (Fig.1, Fig. S1). We selected the L3 vertebra if the L4 vertebra had a grade 2 or 3 fracture by using a semiquantitative method (31).

**Figure 1.**
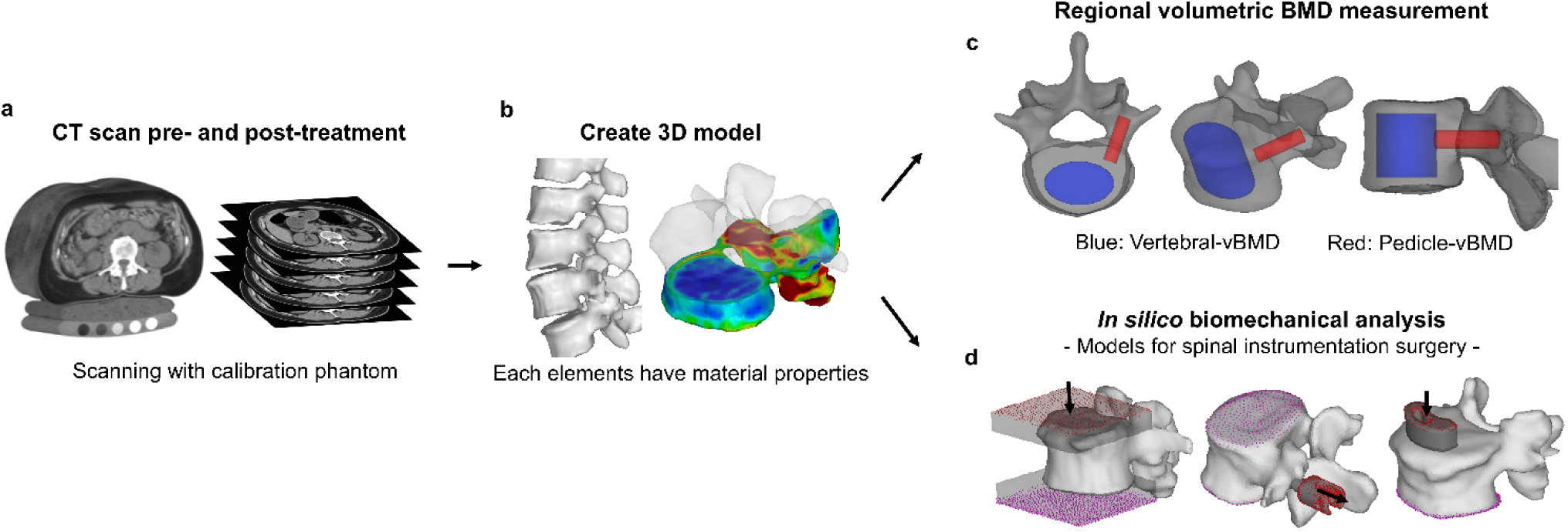
Overview of *in silico* drug assessment for instrumentation surgery. The proposed framework for drug assessment in instrumentation surgery. **a,** CT scans, pre- and post-treatment, with a calibration phantom for quality longitudinal measurements. **b,** Creation of the 3D (dimensional) vertebral models from the CT data. **c,** Regional 3D v-BMD measurements provide accurate BMD assessment. **d,** Biomechanical evaluations of surgical-related parameters using the finite element method.

### Finite Element Methods

Biomechanical parameters related to spinal instrumentation including compression strength (CS), pullout strength of the screw (POS) and cage subsidence strength (CSS) were evaluated by QCT-based FEM using MECHANICAL FINDER. The FEM modeling methods were based on previous studies and are shown in Fig.1 and Movie.S1-4 (18,32). Briefly, finite element models of the L4 vertebrae were constructed from the CT data and examined for the CS (Movie.S2). Then, a pedicle screw and cage were placed according to the spinal fusion surgery so as to evaluate the POS and CSS (18) (Movie.S3, 4). In the CSS model, a banana-shaped PEEK (polyetheretherketone) cage, which was created using Metasequoia 4 (tetraface Inc., Tokyo, Japan), was set 4-mm behind the anterior edge of the upper endplate vertebrae according to the spinal fusion surgery so as to assess the risk of cage subsidence. A compressive displacement was applied to the cage at the cranial end of the vertebrae at ramped displacement increments of 0.02 mm / step. The predicted CCS was identified by a rapid decrease in the force-displacement curve or a rapid increase in the failure elements. The detailed finite element models and materials properties are provided in Table S1.

### Statistical analysis

Fisher’s exact test and the Mann-Whitney U test were used to compare differences between the 2 groups. Dunn’s test was used for multiple comparisons. The correlations between each parameter were determined using Spearman’s rank coefficients. Statistical analyses were performed using Stat Flex Ver. 6 (Artech, Tokyo). All statistical tests were two tailed and results with P-values< 0.05 were considered statistically significant.

## Results

### Patients and baseline demographics

Sixty-six patients in the Romo-group (66 / 69 patients, 95.7%) and 10 patients in the ELD-group (10 / 12 patients, 83.3%) completed the 6 months study follow-up (Romo: 66 / 69 [95.7%], ELD: 10/12 [83.3%], P = ns)(Fig.S1). The reasons for the discontinued study were as follows; loss of motivation (1 patient in Romo, 2 patients in ELD), hospital administration related to vascular event (1 patient in Romo), death unrelated to treatment (1 patient in Romo). Table 1 shows the demographics and baseline characteristics of the groups. Serum TRACP-5b and P1NP were higher in the ELD group, presumably due to a difference in the prior treatment history, but the difference was not significant. BMD measured by DXA and QCT was equivalent in the two groups.

**Table 1.** Baseline patient characteristics. There were no statistical differences between groups for any of the parameters. The data shown are the median (interquartile ranges [IQR]; Q1/Q3) or n (%). The date shown as n or n (%) were analyzed by Fisher’s exact test. The date presented as median IQR were analyzed by Mann- Whitney U test. BMI; bone mass index, SERM; selective estrogen receptor modulator, eGFR; estimated glomerular filtration rate, total-P1PN; total N-terminal propeptide of type 1 procollagen, TRACP-5P; tartrate-resistant acid phosphatase type 5 protein, aBMD; areal bone mineral density, vBMD; volumetric bone mineral density.

| Parameters | All | Romo | ELD |
| --- | --- | --- | --- |
| Age (years) | 76.0 (70.0 / 81.3) | 77.0 (70.0 / 82.3) | 70.0 (75.0 / 66.5) |
| Male (%) | 5 (6.9) | 4 (5.8) | 1 (8.3) |
| BMI | 21.5 (19.6 / 23.4) | 21.3 (19.5 / 23.1) | 23.2 (21.5 / 24.6) |
| Fracture history (%) | 51 (63.0) | 46 (56.8) | 5 (41.7) |
| Smoking (%) | 5 (6.2) | 3 (4.3) | 2 (16.7) |
| Alcohol (%) | 4 (4.9) | 3 (4.3) | 1 (8.3) |
| **Prior treatment** |  |  |  |
| None | 45 (55.6) | 34 (49.3) | 11 (91.7) |
| Bisphosphonate | 11 (13.6) | 11 (15.9) | 0 (0) |
| SERM | 3 (3.7) | 3 (4.3) | 0 (0) |
| Denosumab | 10 (12.3) | 10 (14.5) | 0 (0) |
| Teriparatide | 12 (14.8) | 11 (15.9) | 1 (8.3) |
| **Blood samples** |  |  |  |
| Albumin levels (g / dl) | 4.3 (4.1 / 4.5) | 4.3 (4.1 / 4.5) | 4.4 (4.2 / 4.6) |
| Corrected calcium levels (mg / dl) | 9.4 (9.1 / 9.5) | 9.4 (9.2 / 9.5) | 9.2 (9.0 / 9.5) |
| Phosphorus levels (mg / dl) | 3.6 (3.2 / 4.1) | 3.6 (3.2 / 4.1) | 3.6 (3.5 / 4.0) |
| eGFR (mL / min / 1.73m<sup>2</sup>) | 66.0 (55.0 / 74.0) | 66.0 (54.8 / 74.0) | 68.5 (61.5 / 75.0) |
| **Bone metabolic markers** |  |  |  |
| total-P1NP (µg / ml) | 58.6 (31.3 / 86.2) | 55.0 (29.2 / 81.0) | 74.1 (58.6 / 115.4) |
| TRACP-5b (mU / dL) | 453.0 (287.3 / 571.3) | 428.0 (268.8 / 560.5) | 536.5 (453.0 / 689.0) |
| **Bone mineral density** |  |  |  |
| Spine- aBMD (g / cm<sup>2</sup>) | 0.75 (0.67 / 0.83) | 0.75 (0.66 / 0.83) | 0.77 (0.73 / 0.85) |
| Vertebral- vBMD (mg / cm<sup>3</sup>) | 77.5 (59.5 / 91.7) | 76.8 (57.5 / 91.0) | 79.3 (76.2 / 95.7) |
| Pedicle - vBMD (mg / cm<sup>3</sup>) | 126.1 (93.1 / 145.0) | 122.4 (92.3 / 143.1) | 138.0 (109.4 / 154.8) |

### Safety

The patients observed to undergo adverse events were 21 (30.4%) in the Romo and no patients in the ELD (Fig. S3). Injection-site reactions such as redness, tenderness and swelling were reported by 16 patients (23.2%) in Romo group. These reactions were well recognized at the initial injection (50.0%). Among the patients who had injection-site reactions, five patients (7.3%) experienced these reactions multiple times during the study (Fig. S4). One (1.5%) patient had hypocalcemia and one (1.5%) patient had hypercalcemia in the Romo group, both of which were of mild severity and asymptomatic.

Gastroenteritis was observed in one (1.5%) patient. Two (2.9 %) patients in the Romo group had severe adverse events including one stroke and one death leading to treatment discontinuation, neither of which was thought to be related to the treatment.

### Changes in bone turnover markers showed dual effects of Romosozumab

The changes in the total P1NP and TRACP-5b levels are shown in Fig. S5. In the Romo group, the P1NP level reached its highest value at 3 months (P< 0.001, vs. Baseline), followed by a gradual decrease at 6 months. Percentage changes from baseline of total P1NP was higher in the Romo group compared to the ELD group at 3 and 6 months (all P< 0.001). The P1NP level decreased significantly over the course of the study in the ELD group (3 months; P< 0.01, 6 months; P< 0.01, vs. baseline). The TRACP-5b levels decreased significantly in both groups at 3 and 6 months, and there was no significant difference between groups.

### Greater increases in regional BMD in Romo group

The median percentage changes from baseline in aBMD by DXA at 6 months was 6.61% (Q1/Q3: 2.62 / 12.7) in the Romo group and 0.91% (0.25 / 2.06) in the ELD group (P < 0.001) (Fig. S6a). The 3D-modeling of the vertebrae demonstrates the heterogeneity of the vertebral BMD that is undetectable by DXA measurement. (Fig. 2a). The difference between the groups observed at 6 months was already identifiable in the regional BMD measurement at 3 months (Fig. 2b-e). The Romo group exhibited significantly greater increases in regional BMD including the vertebral body and pedicle at 3 months compared to the ELD group (V-vBMD; Romo vs. ELD: 10.43% [4.39 / 16.77] vs. 1.54% [-4.45 / 2.48], P < 0.001 and P-vBMD; 12.75% [4.94 / 18.08] vs. 2.22% [-0.35 / 5.13], P < 0.001) (Fig. 2f). Remarkably, treatment with romosozumab resulted in noticeable BMD increases not only within the inner pedicle region (corresponding to ROI of the P-vBMD) but also on the cortical surface of the inner pedicle (Fig. 2b, d, e). Similarly, the BMD around the endplate, the corresponding area for cage placement in spinal fusion surgery, increased following romosozumab treatment (Fig. 2d, e). In treatment-naïve patients, while the results were consistent, the group differences were more evident (aBMD: 9.05% [5.13 / 14.4] vs. 0.95% [0.57 / 2.56], P < 0.001, V-vBMD: 12.84% [5.75 / 20.88] vs. 1.33% [-5.37 / 2.11], P < 0.0001 and P-vBMD: 12.75% [4.94 / 17.49] vs. 1.22% [-0.44 / 5.30], P < 0.001,) (Figs. 2g, S6b).

**Figure 2.**
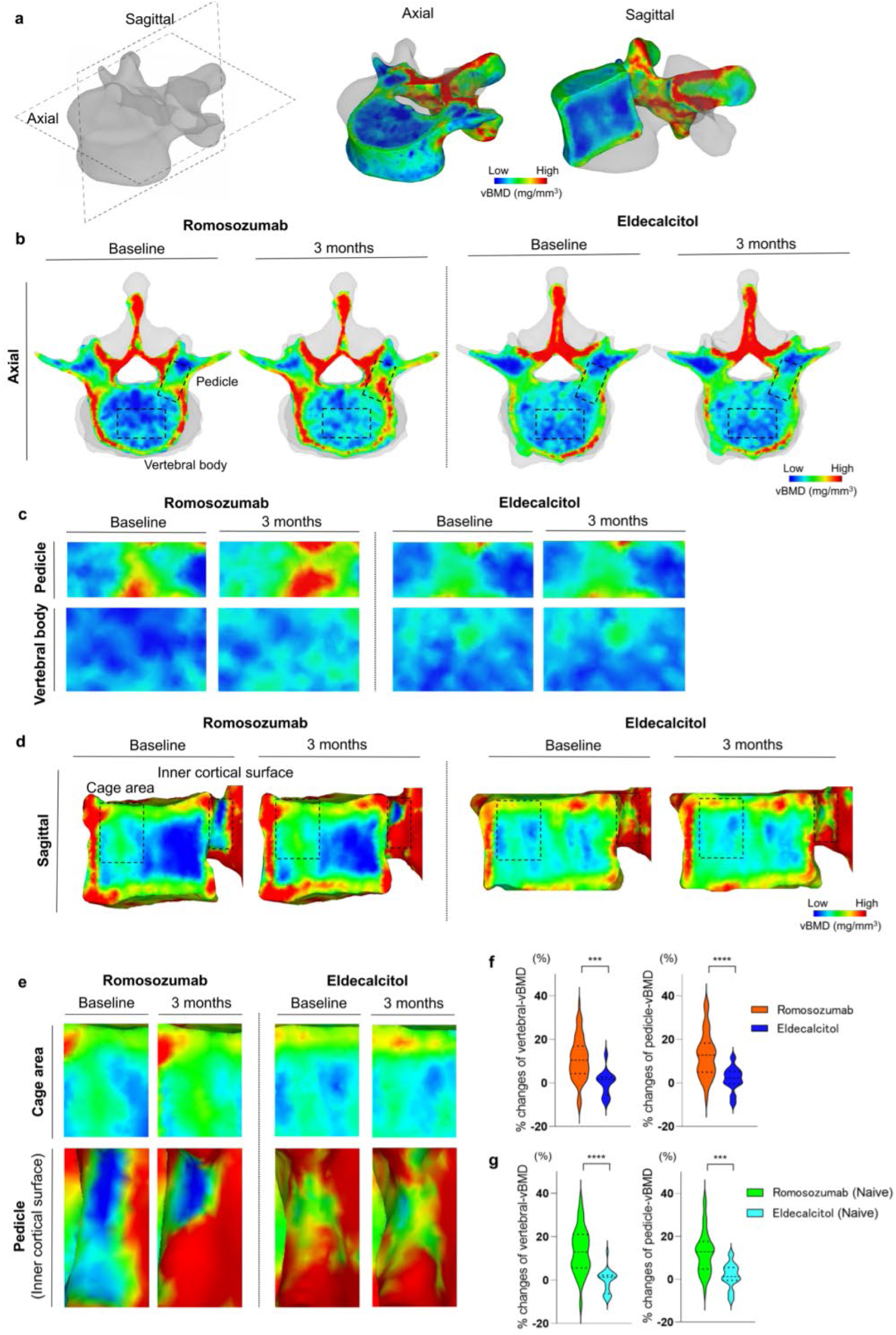
Romosozumab rapidly increases vBMD in the vertebral body and pedicle. **a,** Image illustrating the 3D model of the vertebrae and dimensions in both the axial and sagittal views. Color map representing the distribution of bone mineral density (mg/mm^3^). **b,** Representative axial images of BMD distribution at baseline and 3 months. **c,** Enlarged images of the axial view of the pedicle and vertebral body showing the improvement in BMD with romosozumab treatment in both the pedicle and vertebral body. **d,** Representative sagittal images of BMD at baseline and 3 months. **e,** Enlarged images of the sagittal view of the cage’s corresponding area and inner cortical region of the pedicle, showing the BMD increase with romosozumab. **f. g,** Percentage changes of vertebral-vBMD and pedicle- vBMD between groups following 3 months of treatment. Data are expressed as medians (interquartile ranges). Differences between groups were analyzed using Mann–Whitney U test. *: P < 0.05, **: P < 0.01, ***: P < 0.001, ****: P < 0.0001. vBMD: volumetric bone mineral density

### Significant improvement in biomechanical parameters in the Romo group

Greater gains in the percentage changes from baseline were observed in the Romo group than in the ELD group in all of the biomechanical parameters (compression strength: 11.49% [2.04 / 22.55] vs. 0.74% [-2.85 / 6.89], pullout strength: 20.00% [9.09 / 33.33] vs. 0.00% [0.0 / 10.00], cage subsidence strength: 11.19% [3.08 / 25.31] vs. -0.55% [-6.36 / 4.30], P < 0.01, P < 0.001, P < 0.001, respectively) (Fig. 3d).

**Figure 3.**
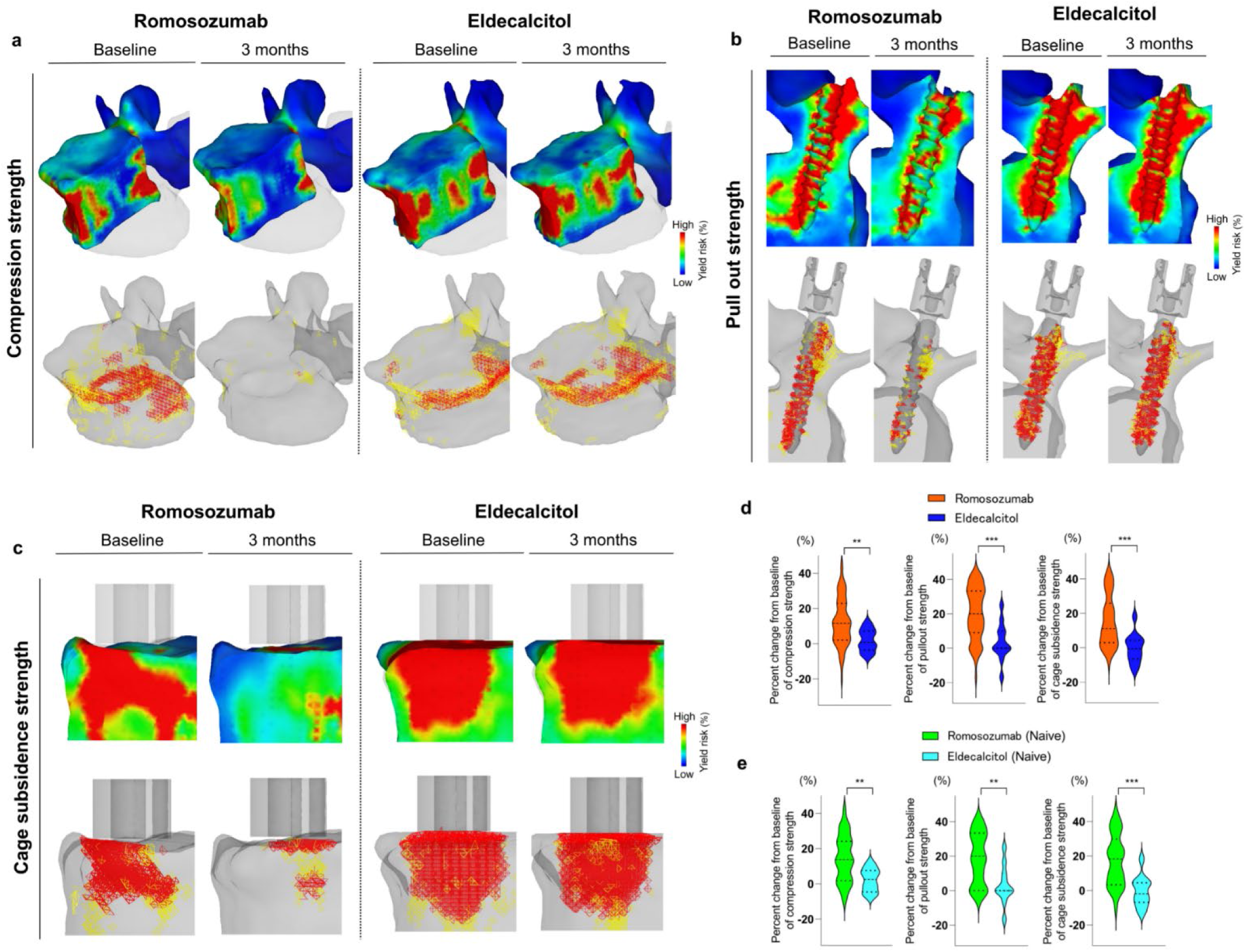
Romosozumab resulted in improvement in all the biomechanical analyses. **a,** Representative images illustrating the compression strength analysis under a 3500 N load, following treatment. The upper images depict the distribution of yield risk (%), while the lower images show the distribution of elements associated with high-risk crushing [yield elements (yellow) and compressive failure elements (red)] both at baseline and 3 months. High yield risk and an increase in crushed elements suggest the potential risk of vertebral fracture. **b,** Representative images showing the pullout strength analysis results when a 200 N force was applied to extract the screw. The upper images depict the distribution of yield risk (%), while the lower images show the distribution of elements associated with high-risk crushing. High yield risk and an increase in crushed elements suggest a potential risk of screw loosening. **c,** Representative images illustrating the cage subsidence strength when a load of 400 N was applied. The upper images depict the distribution of yield risk (%), while the lower images show the distribution of elements associated with high-risk crushing. High yield risk and an increase in crushed elements suggest a potential risk of cage subsidence. **d-e,** Percentage changes in biomechanical parameters between groups following 3 months of treatment. Data are expressed as medians (interquartile ranges). Differences between groups were analyzed using Mann–Whitney U test. *: P < 0.05, **: P < 0.01, ***: P < 0.001

This results were bolstered by the distribution of yield risk and crushed element, which further illustrates the significant improvement following romosozumab treatment (Fig. 3a-c). The findings are consistent among treatment-naïve patients (compression strength: 13.60% [1.81 / 24.1] vs. 2.50% [-3.74 / 7.21], pullout strength: 20.00% [0.00 / 33.33] vs. 0.0% [0.0 / 8.89], cage subsidence strength: 18.27% [3.26 / 28.11] vs. -1.97% [-6.61 / 4.38], P < 0.01, P < 0.01, P < 0.001, respectively) (Fig. 3d). Collectively, these findings suggest a potential contribution of 3 months of romosozumab treatment to the reduction of postoperative complications in instrumentation surgery.

### Regional vBMD had the most significant correlations with biomechanical parameters

Local areal contribution around the implant, as displayed in each biomechanical analysis in Figure 3, led us to examine the correlation between biomechanical parameters with various BMD assessments. The correlations between the BMD and biomechanical parameters are summarized in Table 2. While the aBMD demonstrated mild or no correlations with the biomechanical parameters, the strongest correlations were observed with the regional vBMD across all parameters in both groups.

**Table 2.** The regional BMD is well correlated with the biomechanical parameters. Correlations of the biomechanical parameters and BMD measured by DXA (aBMD) and QCT (vBMD). The biomechanical parameters exhibited significant correlations with each BMD measurement and showed a stronger correlation with the regional vBMD measured by QCT. aBMD: areal bone mineral density, vBMD: volumetric bone mineral density

| Baseline |  | Vertebral strength |  | Pullout strength |  | Cage subsidence strength |  |
| --- | --- | --- | --- | --- | --- | --- | --- |
|  |  | r | p | r | p | r | p |
| All | Spine- aBMD (g / cm<sup>2</sup>) | 0.2640 | 0.0180 | 0.2926 | 0.0080 | 0.1103 | 0.3301 |
|  | Vertebral- vBMD (mg / cm<sup>3</sup>) | 0.6387 | < 0.0001 | 0.5096 | < 0.0001 | 0.6108 | < 0.0001 |
|  | Pedicle - vBMD (mg / cm<sup>3</sup>) | 0.5253 | < 0.0001 | 0.6706 | < 0.001 | 0.3463 | 0.0017 |
| Romo | Spine- aBMD (g / cm<sup>2</sup>) | 0.2959 | 0.0143 | 0.3409 | 0.0042 | 0.1145 | 0.3524 |
|  | Vertebral- vBMD (mg / cm<sup>3</sup>) | 0.6706 | < 0.0001 | 0.4814 | < 0.0001 | 0.5635 | < 0.0001 |
|  | Pedicle - vBMD (mg / cm<sup>3</sup>) | 0.5342 | < 0.0001 | 0.6253 | < 0.0001 | 0.2814 | 0.0201 |
| ELD | Spine- aBMD (g / cm<sup>2</sup>) | 0.1191 | 0.7124 | -0.0424 | 0.8956 | 0.1189 | 0.7129 |
|  | Vertebral- vBMD (mg / cm<sup>3</sup>) | 0.4308 | 0.1621 | 0.1414 | 0.6613 | 0.8531 | 0.0004 |
|  | Pedicle - vBMD (mg / cm<sup>3</sup>) | 0.4308 | 0.1621 | 0.9188 | < 0.0001 | 0.6573 | 0.0202 |
| 3 months |  | Vertebral strength |  | Pullout strength |  | Cage subsidence strength |  |
|  |  | r | p | r | p | r | P |
| All | Vertebral- vBMD (mg / cm<sup>3</sup>) | 0.7106 | < 0.0001 | 0.5470 | < 0.0001 | 0.6294 | < 0.0001 |
|  | Pedicle - vBMD (mg / cm<sup>3</sup>) | 0.5805 | < 0.0001 | 0.6946 | < 0.0001 | 0.3896 | 0.0004 |
| Romo | Vertebral- vBMD (mg / cm<sup>3</sup>) | 0.7474 | < 0.0001 | 0.5324 | < 0.0001 | 0.6177 | < 0.0001 |
|  | Pedicle - vBMD (mg / cm<sup>3</sup>) | 0.5854 | < 0.0001 | 0.6658 | < 0.0001 | 0.3421 | 0.0043 |
| ELD | Vertebral- vBMD (mg / cm<sup>3</sup>) | 0.8392 | 0.0006 | 0.7874 | 0.0024 | 0.8392 | 0.0006 |
|  | Pedicle - vBMD (mg / cm<sup>3</sup>) | 0.4755 | 0.1182 | 0.8647 | 0.0003 | 0.7413 | 0.0006 |

## Discussion

Despite the longstanding recognition through intensive research of osteoporosis as a risk factor for postoperative complications in orthopedic instrumentation surgery, these complications persist as a primary concern and critical priority for surgeons. This challenge is becoming more prominent with the worldwide increase in the aged population (33,34).

One significant limitation of biomechanical implant assessment for surgery, particularly in clinical practice, is the inability to conduct damage or destruction analysis, a method traditionally applied in animal and human cadaver research (35,36). This issue, therefore, presents a difficulty for surgeons when they are trying to determine the effectiveness of osteoporosis treatments in mitigating postoperative complications. Our *in silico* biomechanical analysis, utilizing patient QCT data collected during treatment, offers a broader perspective, and contributes novel insights to the field of bone and implant research.

In this study, romosozumab treatment was associated with larger gains in spine BMD at 6 months, as measured by DXA, consistent with previous trials (24,37,38). The difference observed at 6 months was already evident at 3 months, confirmed by regional vBMD in both the vertebral body and pedicle.

Remarkably, this change began soon after treatment administration, associated with improvements in FEM-estimated biomechanical parameters such as vertebral compression strength, screw pullout strength, and cage subsidence strength. It is noteworthy that these parameters are closely linked with common postoperative complications, suggesting the potential benefits of romosozumab for perioperative management. The notable gains in the both the BMD and FEM-estimated parameters are likely attributed to the dual mechanisms of romosozumab, namely, the enhancement of bone formation and reduction of bone resorption, as demonstrated by the evaluation of bone turnover markers (39–41).

Several strategies have been proposed for managing osteoporosis in patients undergoing spinal surgery, with a broad consensus emphasizing the need for preoperative bone health assessment (11–13). If poor bone health condition is detected, initiation of pharmacological treatment is recommended. Moreover, considering the risk of postoperative complications in patients with osteoporosis, Lubelski and colleagues suggest postponing surgery in order to strengthen the bone prior to the procedure (12). In the light of increases in regional vBMD and potential risk reduction post-surgery shortly after the treatment administration in this study, romosozumab may emerge as a valuable preoperative therapeutic option. This is particularly significant, given that the effectiveness of most osteoporosis therapies is typically not recognized until one or two years after treatment.

Limitations in DXA’s capacity to predict future fractures and postsurgical complications, arising from factors like BMD heterogeneity in the vertebra, osteophyte formation, articular facet hypertrophy, and aortic calcification, are further supported in the present study (5,6,42). We found that all of the biomechanical parameters exhibit their strongest correlations with vBMD around the implant. This implies a significant contribution of region-specific vBMD in postoperative complications. As potential DXA limitations like overestimation of BMD is prevalent in patients with a spinal disorder, evaluation of regional vBMD might yield more clinical utility in assessing the risk of surgery. Furthermore, with the distinct post-treatment changes in regional BMD, along with superior correlations of corresponding regional vBMD with biomechanical parameters, understanding the therapeutic effects of specific regional vBMD could offer potentially valuable insights over traditional DXA assessment for surgeons. However, this hypothesis needs further investigation.

Previous studies on the effects of romosozumab showed increases in both cortical BMD and cortical thickness (25,43). Genant et al. reported that most significant changes in the cortical area predominantly occur in the endocortical region (44). Furthermore, histomorphometry analysis of bone biopsies from a clinical trial showed that the anabolic effect in the initial 2 months of romosozumab treatment mainly arises on the endocortical surface, leading to a 18.3% increase in the mineralizing surface compared to a 4.1% increase in the trabecular bone (26,45). Even though we did not scrutinize parameters related to the cortical and endocortical area due to their ambiguous definition and the limited resolution of imaging studies, we did find that the vBMD in the pedicle responded more favorably to treatment than in the vertebral area. These insights could potentially support previously mentioned studies, given that P- vBMD region of interest presumably includes the endocortical area, whereas V-vBMD mainly comprises trabecular bone.

Consistent with a previous report, romosozumab was generally well tolerated, with no new safety findings observed (24). The most frequently observed adverse events were injection site reactions, which some patients experienced multiple times. While two severe events were noted, the frequency of these events was similar to previous studies (24,38). Although safety concerns, including cardiovascular risk, were raised by romosozumab when compared with alendronate, this risk was not observed in a placebo-controlled trial (24,41,46). In addition, recent studies have shown that romosozumab is not associated with an increased rate of adverse events, regardless of levels of kidney function (47). Nevertheless, careful assessment, including the risk of a cardiovascular event, is desired prior to romosozumab initiation. Moreover, the safety profiles of osteoporosis treatment need to be assessed in the perioperative setting.

This study has several limitations. The first is the non-randomized, open-label design, which was necessitated by the differential effectiveness of each drug in preventing fractures, particularly among our study population, which consisted of patients with relatively severe osteoporosis. Consequently, patient choice followed expert consultation, leading to a disparity in the number of patients in each group. While there was an open-label nature of the treatment assignments, the primary outcomes were measured by investigators who were masked to the group allocation. Another limitation was the small samples size, affecting our power to detect differences between groups. However, the romosozumab groups exhibited significant improvement in primary outcomes, and the sample size was relatively large compared to previous implant-related FEM studies (18,32). In addition, although FEM have been validated in earlier study, our FEM model could not fully mimic the clinical situation (32). Nevertheless, conducting damage or destruction analysis in the clinical setting is inherently impractical. Finally, we were unable to evaluate bone fusion and reoperation rates, one of the endpoints of spinal instrumentation surgery. Nonetheless, both the rigid screw fixation and reduced cage subsidence were potentially facilitated by romosozumab, and thus could beneficially contribute to bone fusion. Further study is warranted to support this notion.

The strengths of this study include the use of a variety of complementary imaging modalities to evaluate the effect of romosozumab on instrumentations surgery, yielding consistent and complementary results. Throughout our novel biomechanical approach, we first established the impact of instrumentation surgery based on the region-specific BMD alternations observed following romosozumab treatment. Such assessments are often a challenge for traditional biomechanical methods, particularly when drugs are involved in the clinical setting. While the present findings highlight a unique benefit of romosozumab for patients with osteoporosis undergoing instrumentation surgery, it is imperative to standardize a multidisciplinary approach, including osteoporosis assessment and treatment, prior to implantation surgery in order to maximize the chance of securing long-term success.

## Data Availability

All data produced in the present work are contained in the manuscript.

## Acknowledgements

We would like to express our sincere gratitude to the staff at the Department of Orthopaedic Surgery at Showa University for their helpful discussion and assistance. We are also grateful to Hideyuki Mimata for the valuable technical assistance. Finally, we thank Ayano Oyamada, Kei Gonsho and Miho Mochizuki for their cooperation in collecting clinical data.

## Supplemental Information

**Supplemental table 1.** Materials properties of each element for finite element models.

| | Young's Modulus<br>(E) (MPa) | Poisson's Ratio | Yield Stress ($\sigma$)<br>(MPa) | Critical Stress<br>(MPa) | Element type | Size/Thickness |
| --- | --- | --- | --- | --- | --- | --- |
| Bone | Keyak (equation 1) | 0.4 | Keyak (equation 2) | St = 0.8Sm | Tetrahedral solid | Less than 2 mm / - |
| Cortical shell | * | 0.4 | Keyak (equation 2) | St = 0.8Sm | Triangular plate | 2 mm / 0.4 mm |
| Cement cap (PMMA) | 2500 | 0.35 | 20.6 | 75.0 | Tetrahedral solid | 2 mm / - |
| Pedicle screw (Titanium alloy) | 108853.8 | 0.28 | 899.3 | 824.7 | Tetrahedral solid | Less than 1 mm / - |
| Cage (PEEK) | 3.6 | 0.3 | 90.0 | 90.0 | Tetrahedral solid | Less than 1 mm / - |

**Supplemental figure 1.**
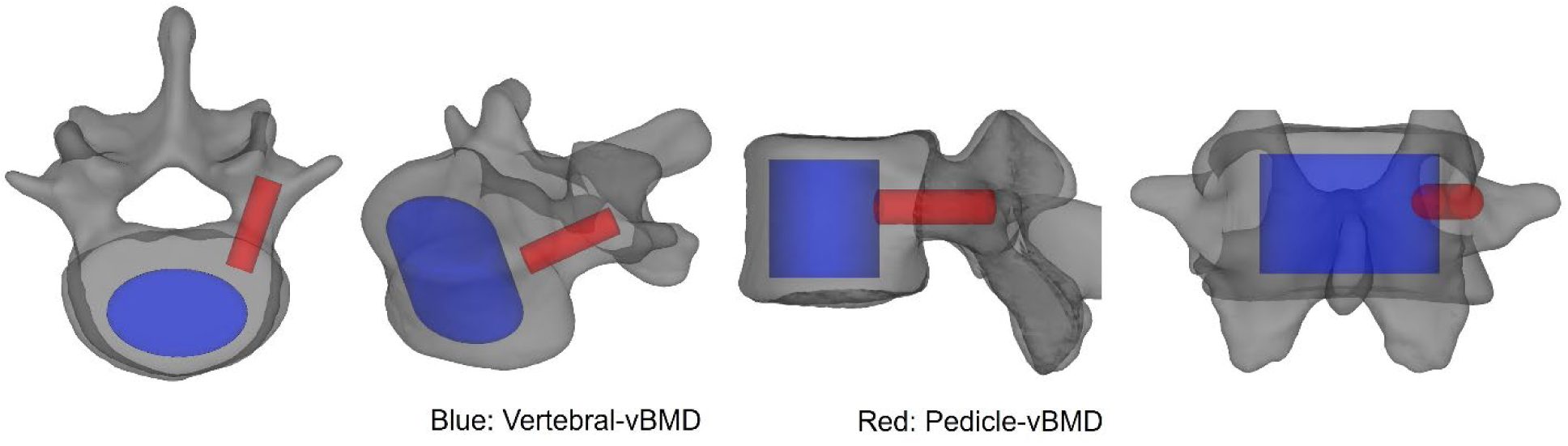
The regions of interest for both pedicle-vBMD and vertebral-vBMD. The red and blue areas illustrate the ROI of the pedicle-vBMD and vertebral-vBMD, respectively. The ROI for the pedicle-vBMD and vertebral-vBMD were manually defined so as to cover the corresponding area, consistent with previous reports (6,18). ROI: region of interest, vBMD: volumetric bone mineral density.

**Supplemental figure 2.**
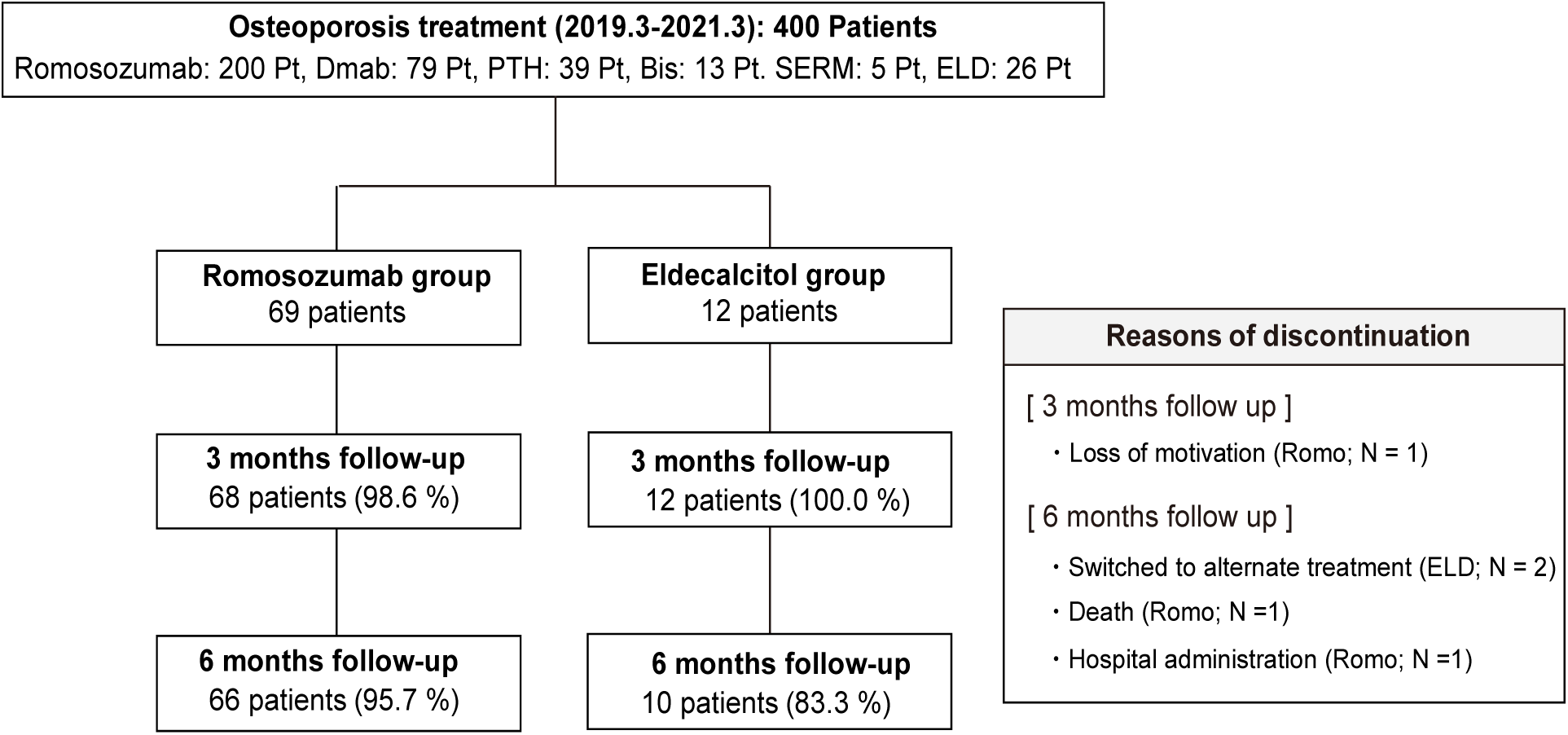
Study diagram. Between March 2019 and March 2021, 400 patients had osteoporosis treatment. 226 patients who were scheduled for romosozumab (Romo) or eldecalcitol (ELD) treatment were assessed for inclusion, with 81 participating in the present study. Patients were divided into two groups based on their treatment (Romo: 69 patients, ELD: 12 patients), and 66 patients (95.7%) from the Romo group and 10 patients (83.3%) from the ELD group completed 6 month follow-up.

**Supplemental figure 3.**
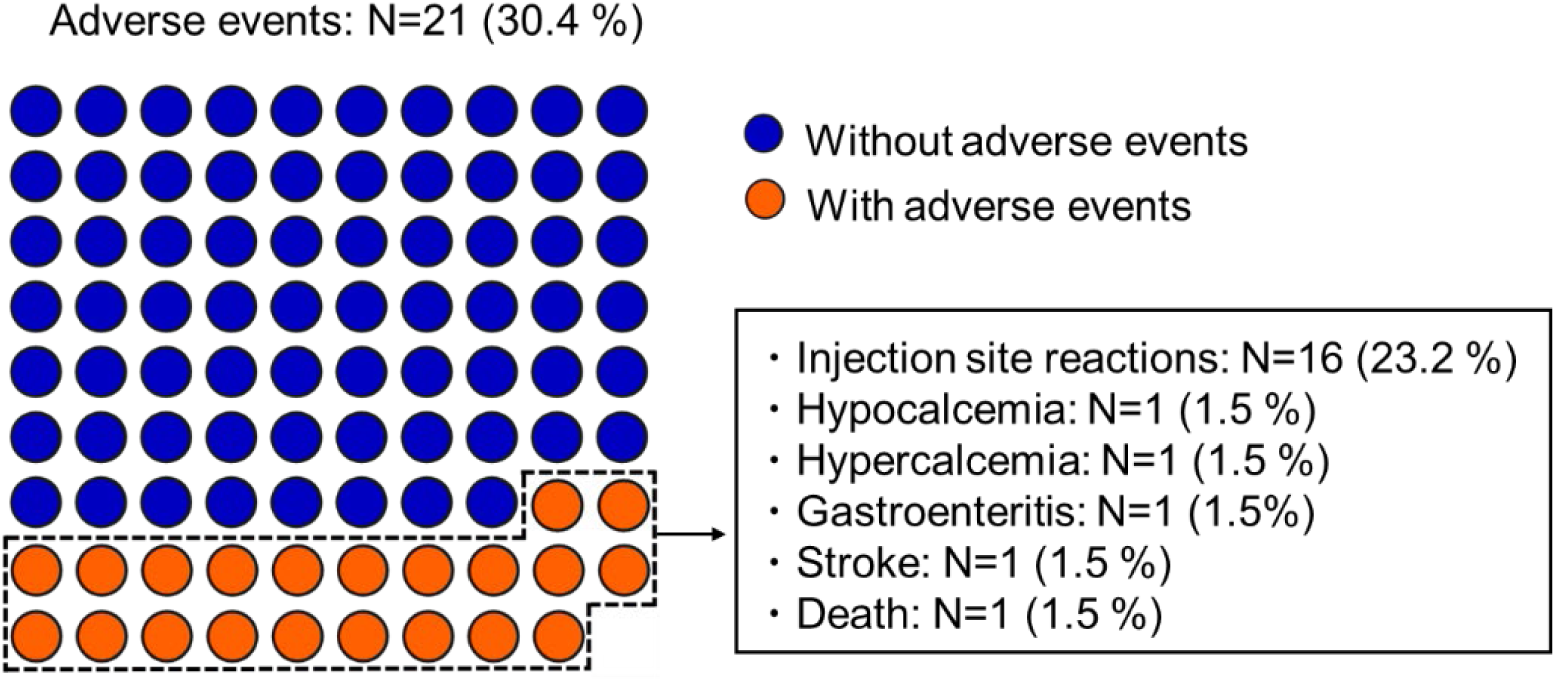
Safety profile of romosozumab treatment for 6 months. The incidence rate of adverse events during 6 months of romosozumab treatment. Blue dots represent patients without any adverse events, whereas the orange dots represent patients who experienced adverse events. Twenty-one patients (30.4%) experienced adverse events following romosozumab treatment. Injection-site reactions were reported by 16 patients (23.2%). Hypocalcemia and hypercalcemia were observed in one patient each (1.5%), respectively. Gastroenteritis was reported by one patient (1.5%). Two (2.9%) patients having severe adverse events included one stroke and one death leading to discontinuation of treatment. These severe adverse events were not thought to be associated with the treatment.

**Supplemental figure 4.**
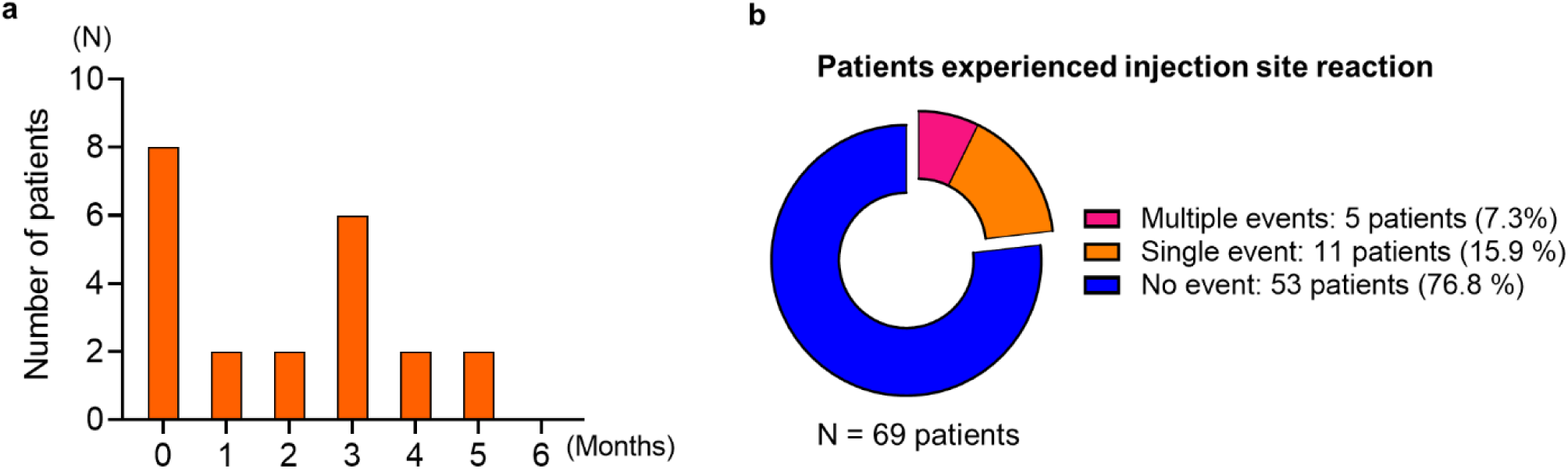
Overview of injection site reactions during romosozumab treatment. **a,** Distribution of injection site reactions. **b,** Proportion of patients experienced multiple injection site reactions. Twenty-two injection-site reactions were reported in 16 patients (23.2%) in the romosozumab group. These reactions were most common at the initial injection (50.0%). Among the patients who had injection-site reactions, five patients (7.3%) experienced these reactions multiple times (2 times: 4 patients, 3 times: 1 patient) during the course of the study.

**Supplemental figure 5.**
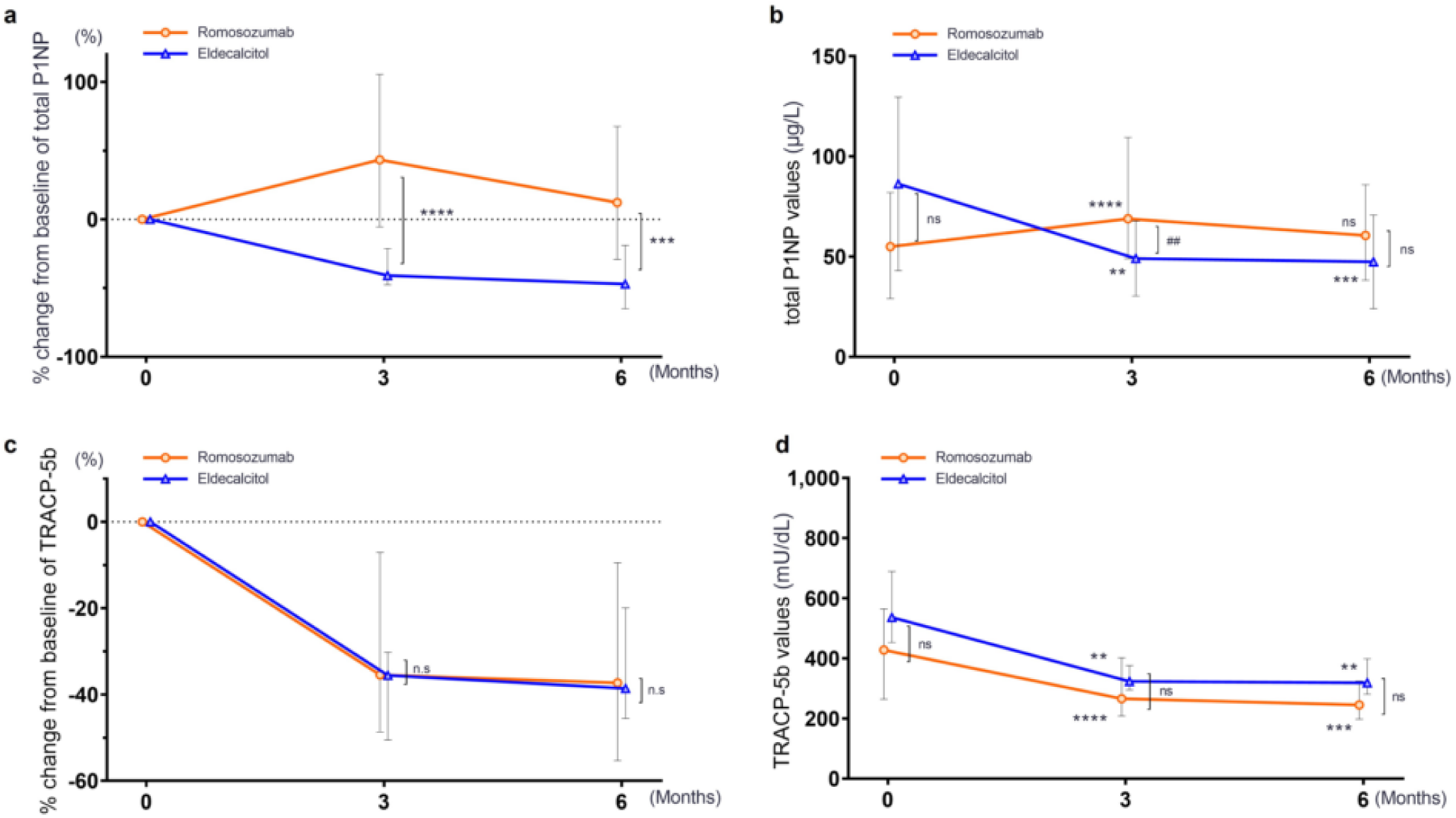
Changes in bone turnover markers during the 6-month treatment. **a,** Percentage change from baseline in total P1NP. **b,** Absolute change from baseline in total P1NP, **c,** Percentage change from baseline in TRACP-5b. **d,** Absolute change from baseline in TRACP-5b. In the Romo group, the P1NP level peaked at 3 months, and the percentage change from baseline of total P1NP was higher compared to the ELD group at both 3 and 6 months. The TRACP-5b levels decreased significantly in both groups, with no significant difference observed between the groups. Data are expressed as medians (interquartile ranges). The difference in the continuous measures across the groups was compared using a one-way analysis by the Dunn test. Differences between groups were analyzed using the Mann–Whitney U test. *: P < 0.05, **: P < 0.01, ***: P < 0.001, ****: P < 0.0001, ns: not significant. total P1NP: total N-terminal propeptide of type 1 procollagen, TRACP-5b: tartrate-resistant acid phosphatase 5b.

**Supplemental figure 6.**
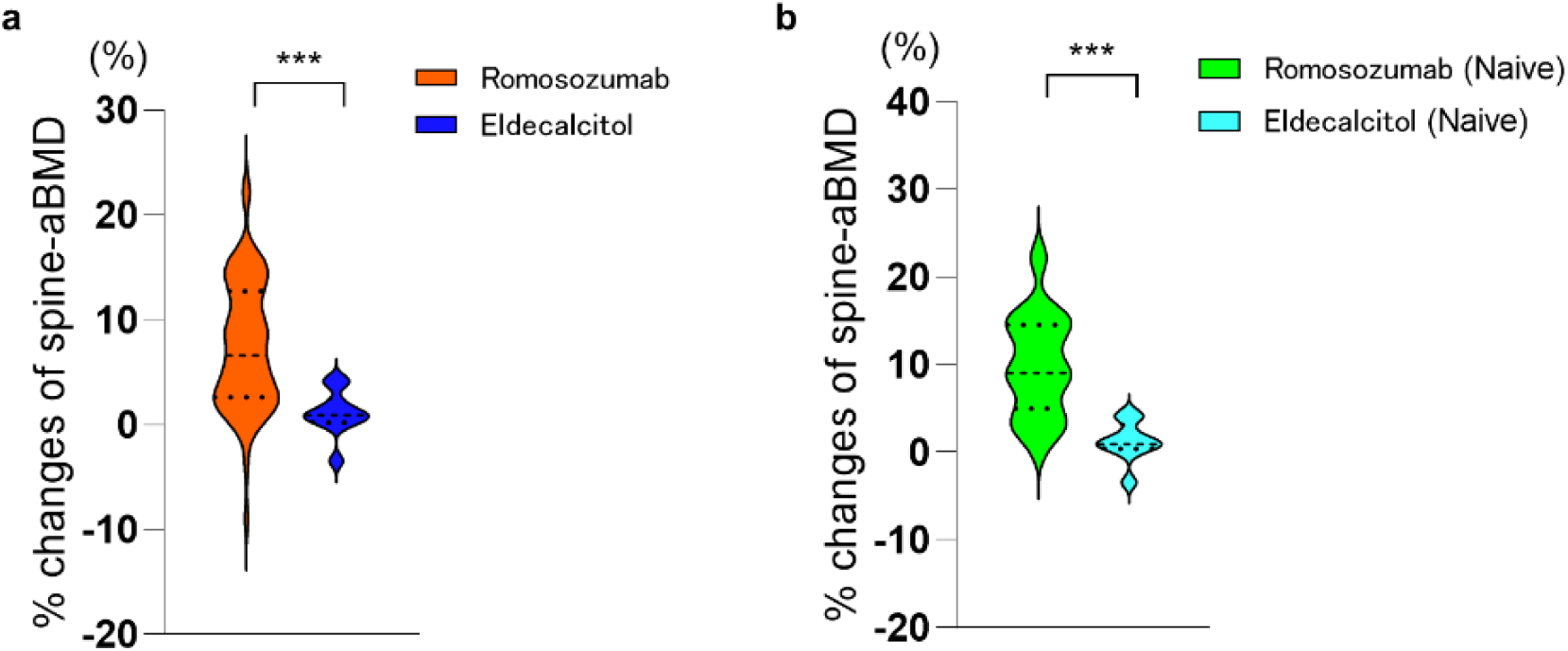
Spine-aBMD assessed by DXA increased significantly following romosozumab treatment at 6 months. Percentage changes of spine-aBMD between groups following 6 months of treatment. **a,** Percentage change in all patients. **b,** Percentage change in patients with treatment naïve. aBMD: areal bone mineral density

**Supplemental Movie1. The modeling methods of a 3D vertebra**

A 3D model of the spine was created based on the CT data, then the L4 vertebra was constructed with 2mm tetrahedral solid elements and 2-mm triangular plates using MECHANICAL FINDER software (version 12.0 extended edition; Research Center of Computational Mechanics, Tokyo, Japan). Each element was assigned individual mechanical properties, and the bone mineral density was integrated to account for bone heterogeneity.

**Supplemental Movie2. Finite element analysis for compression strength**

A compressive displacement was applied to the PMMA (poly methyl-methacrylate) cement cap at the cranial end of the vertebrae, with displacement increments of 0.01 mm / step. The time segment from 0:07-0:17 depicts the distribution of high-risk elements associated with crushing [yield elements (yellow) and compressive failure elements (red)], while 0:18-0:41 illustrates the distribution of yield risk. High yield risk and an increase in compressive failure elements suggest a potential risk for compression fracture.

**Supplemental Movie3. Finite element analysis for pullout strength**

A screw was implanted according to the conventional pedicle screw trajectory. The vertebra was fully fixed in all directions, and an incremental tensile loading rate of 20 Newton (N) / step was applied to the screw head. From 0:06-0:17 the images show the distribution of high-risk elements associated with crushing [yield elements (yellow) and compressive failure elements (red)], while 0:18-0:30 illustrates the distribution of yield risk. High yield risk and an increase in compressive failure elements suggest a potential risk for compression fracture.

**Supplemental Movie4. Finite element analysis for cage subsidence strength**

A PEEK (polyetheretherketone) cage was positioned 5-mm behind the anterior edge of the upper endplate vertebrae to assess the risk of cage subsidence. A compressive displacement was applied to the PEEK cage at the cranial end of the vertebrae, using ramped displacement increments of 0.01 mm / step. The time segment of the 0:05-0:15 images shows the distribution of high-risk elements associated with crushing [yield elements (yellow) and compressive failure elements (red)], while 0:20-0:30 illustrates the distribution of yield risk. High yield risk and an increase in compressive failure elements suggest a potential risk of cage subsidence.

## References

1. Weiser TG, Haynes AB, Molina G, Lipsitz SR, Esquivel MM, Uribe-Leitz T, et al. Estimate of the global volume of surgery in 2012: an assessment supporting improved health outcomes. Lancet. 2015;385:S11.

2. James SL, Abate D, Abate KH, Abay SM, Abbafati C, Abbasi N, et al. Global, regional, and national incidence, prevalence, and years lived with disability for 354 Diseases and Injuries for 195 countries and territories, 1990-2017: A systematic analysis for the Global Burden of Disease Study 2017. Lancet. 2018;392(10159):1789–858.

3. Etzioni DA, Liu JH, Maggard MA, Ko CY. The Aging Population and Its Impact on the Surgery Workforce. Ann Surg. 2003;238(2):170–7.

4. Punnoose A, Claydon-Mueller LS, Weiss O, Zhang J, Rushton A, Khanduja V. Prehabilitation for Patients Undergoing Orthopedic Surgery: A Systematic Review and Meta-analysis. JAMA Netw Open. 2023;6(4):E238050.

5. Keaveny TM, Adams AL, Fischer H, Brara HS, Burch S, Guppy KH, et al. Increased risks of vertebral fracture and reoperation in primary spinal fusion patients who test positive for osteoporosis by Biomechanical Computed Tomography analysis. Spine J [Internet]. 2023;23(3):412–24. Available from: 10.1016/j.spinee.2022.10.018

6. Ishikawa K, Toyone T, Shirahata T, Kudo Y, Matsuoka A, Maruyama H, et al. A Novel Method for the Prediction of the Pedicle Screw Stability. Clin Spine Surg. 2018;31(9):E473–80.

7. Bliuc D, Nguyen ND, Milch VE, Nguyen T V., Eisman JA, Center JR. Mortality risk associated with low-trauma osteoporotic fracture and subsequent fracture in men and women. JAMA. 2009;301(5):513–21.

8. Center JR, Bliuc D, Nguyen ND, Nguyen T V., Eisman JA. Osteoporosis medication and reduced mortality risk in elderly women and men. J Clin Endocrinol Metab. 2011;96(4):1006–14.

9. Abrahamsen B, Osmond C, Cooper C. Osteoporosis: Treat or let die twice more likely. J Bone Miner Res. 2015;30(9):1553–9.

10. Miller AN, Lake AF, Emory CL. Establishing a fracture liaison service: An orthopaedic approach. J Bone Jt Surg - Am Vol. 2015;97(8):675–81.

11. Sardar ZM, Coury JR, Cerpa M, Dewald CJ, Ames CP, Shuhart C, et al. Best Practice Guidelines for Assessment and Management of Osteoporosis in Adult Patients Undergoing Elective Spinal Reconstruction. Spine (Phila Pa 1976). 2022;47(2):128–35.

12. Lubelski D, Choma TJ, Steinmetz MP, Harrop JS, Mroz TE. Perioperative medical management of spine surgery patients with osteoporosis. Neurosurgery. 2015;77(4):S92–7.

13. Zhang A, Khatri S, Balmaceno-Criss M, Alsoof D, Daniels AH. Medical optimization of osteoporosis for adult spinal deformity surgery: a state-of-the-art evidence-based review of current pharmacotherapy. Spine Deform [Internet]. 2022;11(3):579–96. Available from: 10.1007/s43390-022-00621-6

14. Chang HK, Ku J, Ku J, Kuo YH, Chang CC, Wu CL, et al. Correlation of bone density to screw loosening in dynamic stabilization: an analysis of 176 patients. Sci Rep [Internet]. 2021;11(1):1–7. Available from: 10.1038/s41598-021-95232-y

15. Bredow J, Boese CK, Werner CML, Siewe J, Löhrer L, Zarghooni K, et al. Predictive validity of preoperative CT scans and the risk of pedicle screw loosening in spinal surgery. Arch Orthop Trauma Surg. 2016;136(8):1063–7.

16. Pearson HB, Dobbs CJ, Grantham E, Niebur GL, Chappuis JL, Boerckel JD. Intraoperative biomechanics of lumbar pedicle screw loosening following successful arthrodesis. J Orthop Res. 2017;35(12):2673–81.

17. Kaiser J, Allaire B, Fein PM, Lu D, Adams A, Kiel DP, et al. Heterogeneity and Spatial Distribution of Intravertebral Trabecular Bone Mineral Density in the Lumbar Spine Is Associated With Prevalent Vertebral Fracture. J Bone Miner Res. 2020;35(4):641–8.

18. Tani S, Ishikawa K, Kudo Y, Tsuchiya K, Matsuoka A, Maruyama H, et al. The effect of denosumab on pedicle screw fixation: a prospective 2-year longitudinal study using finite element analysis. J Orthop Surg Res. 2021;16(1):1–10.

19. Parisien A, Wai EK, Elsayed MSA, Frei H. Subsidence of Spinal Fusion Cages: A Systematic Review. Int J Spine Surg. 2022;16(6):1103–18.

20. Zhang Y, Jiang Y, Zou D, Yuan B, Ke HZ, Li W. Therapeutics for enhancement of spinal fusion: A mini review. J Orthop Transl. 2021 Nov 1;31:73–9.

21. Ebata S, Takahashi J, Hasegawa T, Mukaiyama K, Isogai Y, Ohba T, et al. Role of weekly teriparatide administration in osseous union enhancement within six months after posterior or transforaminal lumbar interbody fusion for osteoporosis-associated lumbar degenerative disorders: A multicenter, prospective randomized study. J Bone Jt Surg - Am Vol [Internet]. 2017 [cited 2022 May 28];99(5):365–72. Available from: https://journals.lww.com/jbjsjournal/Fulltext/2017/03010/Role_of_Weekly_Teriparatide_Administration_in.1.aspx

22. Yagi M, Ohne H, Konomi T, Fujiyoshi K, Kaneko S, Komiyama T, et al. Teriparatide improves volumetric bone mineral density and fine bone structure in the UIV+1 vertebra, and reduces bone failure type PJK after surgery for adult spinal deformity. Osteoporos Int [Internet]. 2016;27(12):3495–502. Available from: 10.1007/s00198-016-3676-6

23. Ohtori S, Inoue G, Orita S, Yamauchi K, Eguchi Y, Ochiai N, et al. Comparison of teriparatide and bisphosphonate treatment to reduce pedicle screw loosening after lumbar spinal fusion surgery in postmenopausal women with osteoporosis from a bone quality perspective. Spine (Phila Pa 1976). 2013;38(8):487–92.

24. Cosman F, Crittenden DB, Adachi JD, Binkley N, Czerwinski E, Ferrari S, et al. Romosozumab Treatment in Postmenopausal Women with Osteoporosis. N Engl J Med. 2016;375(16):1532–43.

25. Graeff C, Campbell GM, Peña J, Borggrefe J, Padhi D, Kaufman A, et al. Administration of romosozumab improves vertebral trabecular and cortical bone as assessed with quantitative computed tomography and finite element analysis. Bone [Internet]. 2015;81:364–9. Available from: 10.1016/j.bone.2015.07.036

26. Chavassieux P, Chapurlat R, Portero-Muzy N, Roux JP, Garcia P, Brown JP, et al. Bone-Forming and Antiresorptive Effects of Romosozumab in Postmenopausal Women With Osteoporosis: Bone Histomorphometry and Microcomputed Tomography Analysis After 2 and 12 Months of Treatment. J Bone Miner Res. 2019;34(9):1597–608.

27. Orimo H, Nakamura T, Hosoi T, Iki M, Uenishi K, Endo N, et al. Japanese 2011 guidelines for prevention and treatment of osteoporosis-executive summary. Arch Osteoporos. 2012;7(1–2):3–20.

28. Ishikawa K, Nagai T, Sakamoto K, Ohara K, Eguro T, Ito H, et al. High bone turnover elevates the risk of denosumab-induced hypocalcemia in women with postmenopausal osteoporosis. Ther Clin Risk Manag. 2016;12:1831–40.

29. Tsuchiya K, Ishikawa K, Kudo Y, Tani S, Nagai T, Toyone T, et al. Analysis of the subsequent treatment of osteoporosis by transitioning from bisphosphonates to denosumab, using quantitative computed tomography: A prospective cohort study. Bone Reports [Internet]. 2021;14(April):101090. Available from: 10.1016/j.bonr.2021.101090

30. Kuroda T, Ishikawa K, Nagai T, Fukui T, Hirano T, Inagaki K. Quadrant Analysis of Quantitative Computed Tomography Scans of the Femoral Neck Reveals Superior Region-Specific Weakness in Young and Middle-Aged Men With Type 1 Diabetes Mellitus. J Clin Densitom [Internet]. 2018;21(2):172–8. Available from: 10.1016/j.jocd.2017.01.005

31. Harry K Genant, Chun Y Wu, Corner Van Kuijk, Michael C Nevitt. Using a Semiquantitative Technique. J Bone Miner Res. 1993;8(9):1137–48.

32. Matsuura Y, Giambini H, Ogawa Y, Fang Z, Thoreson AR, Yaszemski MJ, et al. Specimen- specific nonlinear finite element modeling to predict vertebrae fracture loads after vertebroplasty. Spine (Phila Pa 1976). 2014;39(22):E1291–6.

33. Arsen M Pankovich, Imad E Tarabishy. Remote nailing of intertrochanteric and subtrochanteric fractures of the femur. Instr Course Lect. 1983;32(4):303–16.

34. Laros GS. The role of osteoporosis in intertrochanteric fractures. Orthop Clin North Am [Internet]. 1980;11(3):525–37. Available from: 10.1016/S0030-5898(20)31455-3

35. Keiler A, Schmoelz W, Erhart S, Gnanalingham K. Primary stiffness of a modified transforaminal lumbar interbody fusion cage with integrated screw fixation: Cadaveric biomechanical study. Spine (Phila Pa 1976). 2014;39(17).

36. Fu X, Tan J, Sun CG, Leng HJ, Xu YS, Song CL. Intraosseous injection of simvastatin in poloxamer 407 hydrogel improves pedicle-screw fixation in ovariectomized minipigs. J Bone Jt Surg - Am Vol. 2016;98(22):1924–32.

37. Brown JP, Engelke K, Keaveny TM, Chines A, Chapurlat R, Foldes AJ, et al. Romosozumab improves lumbar spine bone mass and bone strength parameters relative to alendronate in postmenopausal women: results from the Active-Controlled Fracture Study in Postmenopausal Women With Osteoporosis at High Risk (ARCH) trial. J Bone Miner Res. 2021;36(11):2139–52.

38. Langdahl BL, Libanati C, Crittenden DB, Bolognese MA, Brown JP, Daizadeh NS, et al. Romosozumab (sclerostin monoclonal antibody) versus teriparatide in postmenopausal women with osteoporosis transitioning from oral bisphosphonate therapy: a randomised, open-label, phase 3 trial. Lancet [Internet]. 2017;390(10102):1585–94. Available from: 10.1016/S0140-6736(17)31613-6

39. Khosla S, Hofbauer LC. Osteoporosis treatment: recent developments and ongoing challenges. lancet Diabetes Endocrinol. 2017 Nov;5(11):898–907.

40. Xu H, Wang W, Liu X, Huang W, Zhu C, Xu Y, et al. Targeting strategies for bone diseases: signaling pathways and clinical studies. Signal Transduct Target Ther [Internet]. 2023;8(1):202. Available from: http://www.ncbi.nlm.nih.gov/pubmed/37198232%0Ahttp://www.pubmedcentral.nih.gov/articlerender.fcgi?artid=PMC10192458

41. Saag KG, Petersen J, Brandi ML, Karaplis AC, Lorentzon M, Thomas T, et al. Romosozumab or Alendronate for Fracture Prevention in Women with Osteoporosis. N Engl J Med. 2017;377(15):1417–27.

42. Rand T, Seidl G, Kainberger F, Resch A, Hittmair K, Schneider B, et al. Impact of spinal degenerative changes on the evaluation of bone mineral density with dual energy X-ray absorptiometry (DXA). Calcif Tissue Int. 1997;60(5):430–3.

43. Poole KES, Treece GM, Pearson RA, Gee AH, Bolognese MA, Brown JP, et al. Romosozumab Enhances Vertebral Bone Structure in Women With Low Bone Density. J Bone Miner Res. 2022;37(2):256–64.

44. Genant HK, Engelke K, Bolognese MA, Mautalen C, Brown JP, Recknor C, et al. Effects of Romosozumab Compared With Teriparatide on Bone Density and Mass at the Spine and Hip in Postmenopausal Women With Low Bone Mass. J Bone Miner Res. 2017;32(1):181–7.

45. Eriksen EF, Chapurlat R, Boyce RW, Shi Y, Brown JP, Horlait S, et al. Modeling-Based Bone Formation After 2 Months of Romosozumab Treatment: Results From the FRAME Clinical Trial. J Bone Miner Res. 2022;37(1):36–40.

46. Ayers C, Kansagara D, Lazur B, Fu R, Kwon A, Harrod C. Effectiveness and Safety of Treatments to Prevent Fractures in People With Low Bone Mass or Primary Osteoporosis: A Living Systematic Review and Network Meta-analysis for the American College of Physicians. Ann Intern Med. 2023;176(2):182–95.

47. Miller PD, Adachi JD, Albergaria BH, Cheung AM, Chines AA, Gielen E, et al. Efficacy and Safety of Romosozumab Among Postmenopausal Women With Osteoporosis and Mild-to- Moderate Chronic Kidney Disease. J Bone Miner Res. 2022;37(8):1437–45.

